# Prediction of Suicidal Ideation in the Canadian Community *Health Survey - Mental Health Component* Using Deep Learning

**DOI:** 10.1101/19010413

**Authors:** Sneha Desai, Myriam Tanguay-Sela, David Benrimoh, Robert Fratila, Eleanor Brown, Kelly Perlman, Ann John, Marcos DelPozo-Banos, Nancy Low, Sonia Israel, Lisa Palladini, Gustavo Turecki

## Abstract

**Introduction:** Suicidal ideation (SI) is prevalent in the general population, and is a prominent risk factor for suicide. However, predicting which patients are likely to have SI remains a challenge. Deep Learning (DL) may be a useful tool in this context, as it can be used to find patterns in complex, heterogeneous, and incomplete psychiatric datasets. An automated screening system for SI could help prompt clinicians to be more attentive to patients at risk for suicide.

**Methods:** Using the Canadian Community Health Survey - Mental Health Component, we trained a DL model based on 23,859 survey responses to predict lifetime SI on an individual patient basis. Models were created to predict both lifetime and last 12 month SI. We reduced 582 possible model parameters captured by the survey to 96 and 21 feature versions of the models. Models were trained using an undersampling procedure that balanced the training set between SI and non-SI respondents; validation was done on held-out data.

**Results:** AUC was used as the main model metric. For lifetime SI, the 96 feature model had an AUC of 0.79 and the 21 feature model had an AUC of 0.75. For SI in the last 12 months the 96 feature model had an AUC of 0.76 and the 21 feature model had an AUC of 0.69. DL outperformed random forest classifiers.

**Discussion:** Although requiring further study to ensure clinical relevance and sample generalizability, this study is a proof-of-concept for the use of DL to improve prediction of SI. This kind of model would help start conversations with patients which could lead to improved care and, it is hoped, a reduction in suicidal behavior.

## 1. Introduction

Suicide is one of the leading causes of death across the world, accounting for approximately 800,000 deaths each year with the number of attempts an order of magnitude higher (World Health Organization [WHO], 2018). Globally, suicide accounts for 16% of injury deaths (World Health Organization [WHO], 2012) and is the second leading cause of death in young people aged 15 to 29 years (World Health Organization [WHO], 2014). This makes suicide prevention a major public health concern (Turecki & Brent, 2016). According to a meta-analysis of 365 studies, among the most important risk factors for suicide attempts and deaths are previous self-injurious behaviors and suicidal ideation (Franklin et al., 2017). Suicidal ideation includes any thoughts about suicide such as a desire for or planning of a suicide attempt and must be distinguished from actual suicidal attempts which involve acting on these thoughts (Beck, Kovacs, & Weissman, 1979). This is addressed by item 9 of the depression module of the Patient Health Questionnaire (PHQ-9) as “thoughts that you would be better off dead or of hurting yourself in some way” (Kroenke & Spitzer, 2002). Importantly, there is a moderately strong association between suicide and suicidal ideation, making it an important factor to consider when assessing suicide risk (McHugh et al., 2019; Hubers et al., 2016). It is important to note that this association is heterogeneous and has low positive predictive value and sensitivity (McHugh et al., 2019). As such, it is clear that not all patients who later die by suicide will express suicidal ideation. On the other hand, suicidal ideation is much more common than attempts, and many patients who express suicidal ideation do not actually attempt suicide (Srivastava & Kumar, 2005). Regardless, proactive detection of ideation is helpful in the identification of patients at risk of suicide.

In current clinical practice, the primary method for identifying the presence of suicidal ideation is through direct questioning or patient self-report. Suicidal ideation can be also be identified and characterized using the instruments, such as the PHQ-9 or the Scale for Suicide Ideation. This method is limited because patients may conceal suicidal intentions from clinicians, and additionally, clinicians often fail to even ask about suicidal ideation (Bongiovi-Garcia et al., 2009). It would therefore be clinically useful to identify which patients may be at risk of suicidal ideation without needing to ask them directly, perhaps by using an automated screening system incorporated into the electronic medical record, as this would allow clinicians to identify patients who might benefit from further assessment and resources.

In the current literature, the vast majority of studies focus on identifying individual predictors or an interaction of only a few factors, resulting in small effect sizes with low predictive value (Franklin et al., 2017). As such, it may be useful to employ more sophisticated methods that can consider a large number of factors when making predictions. Machine learning, which allows for the creation of models that can consider many factors and identify complex relationships between them, may be an ideal tool for identifying people with suicidal ideation. While a few machine learning models have been created to predict suicide attempts (DelPozo-Banos et al., 2018, Passos et al., 2016, Walsh et al., 2017), we found only one that aimed at predicting suicidal ideation (Jordan et al., 2018). This study investigated suicidal ideation in a primary care patient sample, as a significant number of people who die by suicide have contact with primary healthcare providers in the month and year prior to their suicide (45% and 75%, respectively) (Turecki & Brent, 2016; Jordan et al., 2018). Jordan and colleagues’ model found that assessing four of the PHQ-9 items was sufficient to predict the presence of suicidal ideation.

Our objective was to train a model to predict suicidal ideation in the general population, thus broadening the scope of application by including potential suicide victims who would not seek medical attention prior to their suicide attempt or who have infrequent contact with clinicians. With this goal, we chose to use a deep learning model for a number of reasons. Firstly, deep learning models can be robust to missing data (Cai et al., 2018), which is common in clinical datasets. More importantly, these models are designed to find complex, non-linear patterns in data without requiring us to specify mediators or moderators, allowing us to better approximate the intricate relationships between the multitude of variables that put an individual at risk for suicidal thoughts.

Ideally, our prediction model would be paired with a clinical decision support system (CDSS) that alerts clinicians and other healthcare practitioners to patients who may require further assessment and monitoring of possible suicidal thoughts. Such a tool would connect patients with their clinicians, allowing patients to fill out requested questionnaires and track their progress, while providing clinicians with an organized interface to follow each of their patients and their individual profiles. Similar tools have been found to be clinically useful in detecting and reducing sepsis mortality, and predicting oral cancer recurrence (Exarchos, Goletsis, & Fotiadis, 2012; Manaktala & Claypool, 2017).

Additionally, we hoped to use our machine learning approach to elucidate which patient characteristics are involved in determining the risk for suicidal ideation. This is important not only from the clinical perspective - that is, for understanding the factors that might cause suicidal ideation in an individual person - but also from the public health perspective, as we may discover risk factors for suicidal ideation amenable to intervention via social programs.

## 2. Methods

### 2.1 Dataset

The *Canadian Community Health Survey - Mental Health Component* data was collected in 2012 cross-sectionally for 25,113 people of ages 15 and over living in the ten provinces of Canada. The data was collected either by telephone or in person and 582 data points were collected per respondent. Participants were asked about the presence of suicidal ideation in their lifetime and in the last twelve months. We attempted to predict participant answers to each of these questions separately. We included only subjects who gave a firm “yes” or “no” to the questions about suicidal ideation to maximize the discriminative ability of our model. Other responses were “not applicable”, “don’t know”, “refusal” and “not stated”. This reduced our sample size for the prediction of lifetime suicidal ideation to 23,859 with 21,597 responding “no” and 2,262 responding “yes” and the sample size for the 12 months suicidal ideation prediction to 3,441 with 2,512 responding “yes” and 929 responding “no”. The size and makeup of both these subsets of the data are summarized in Table 7. There were 485 people who responded “yes” to both questions.

**Table 1.**
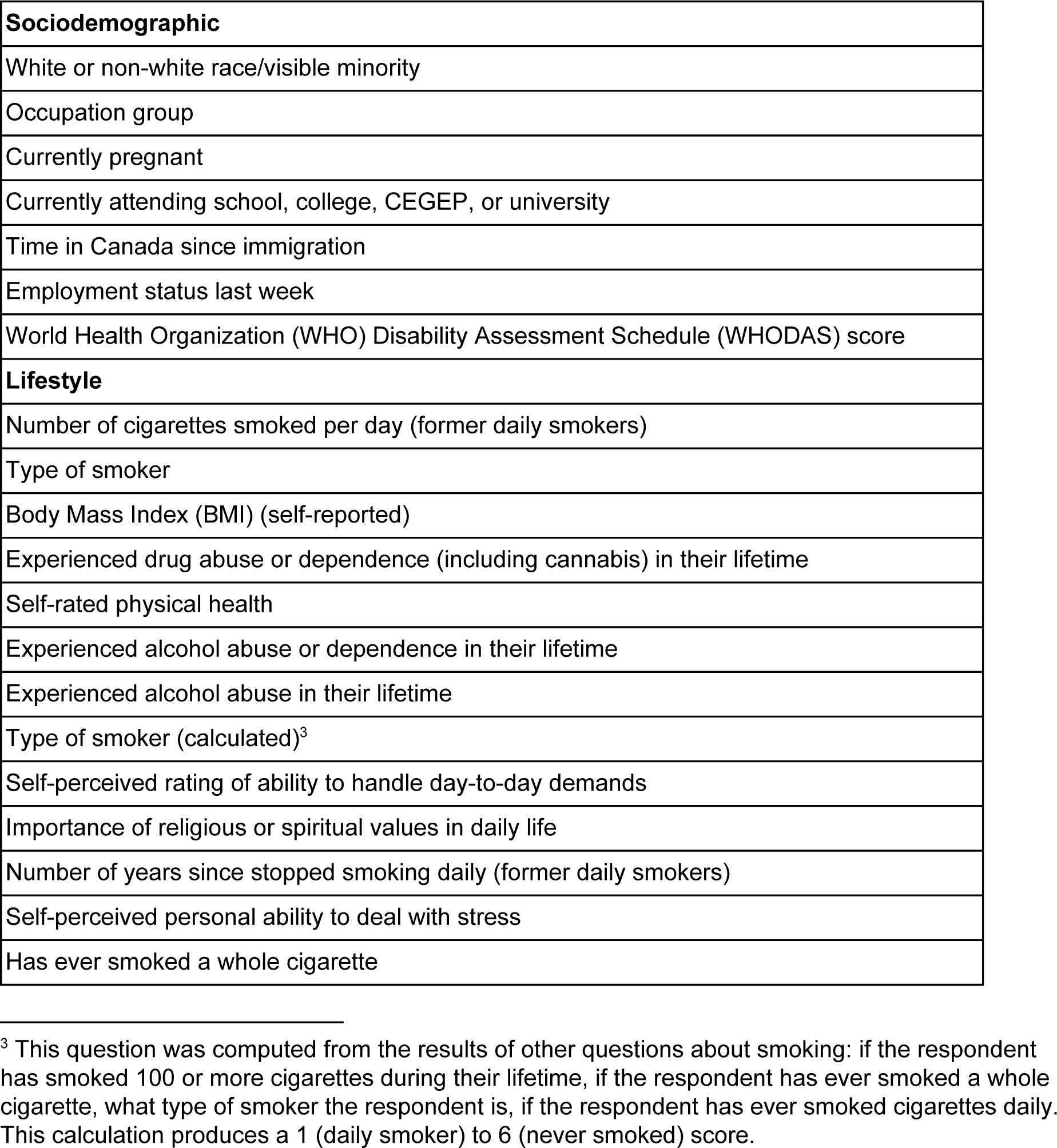
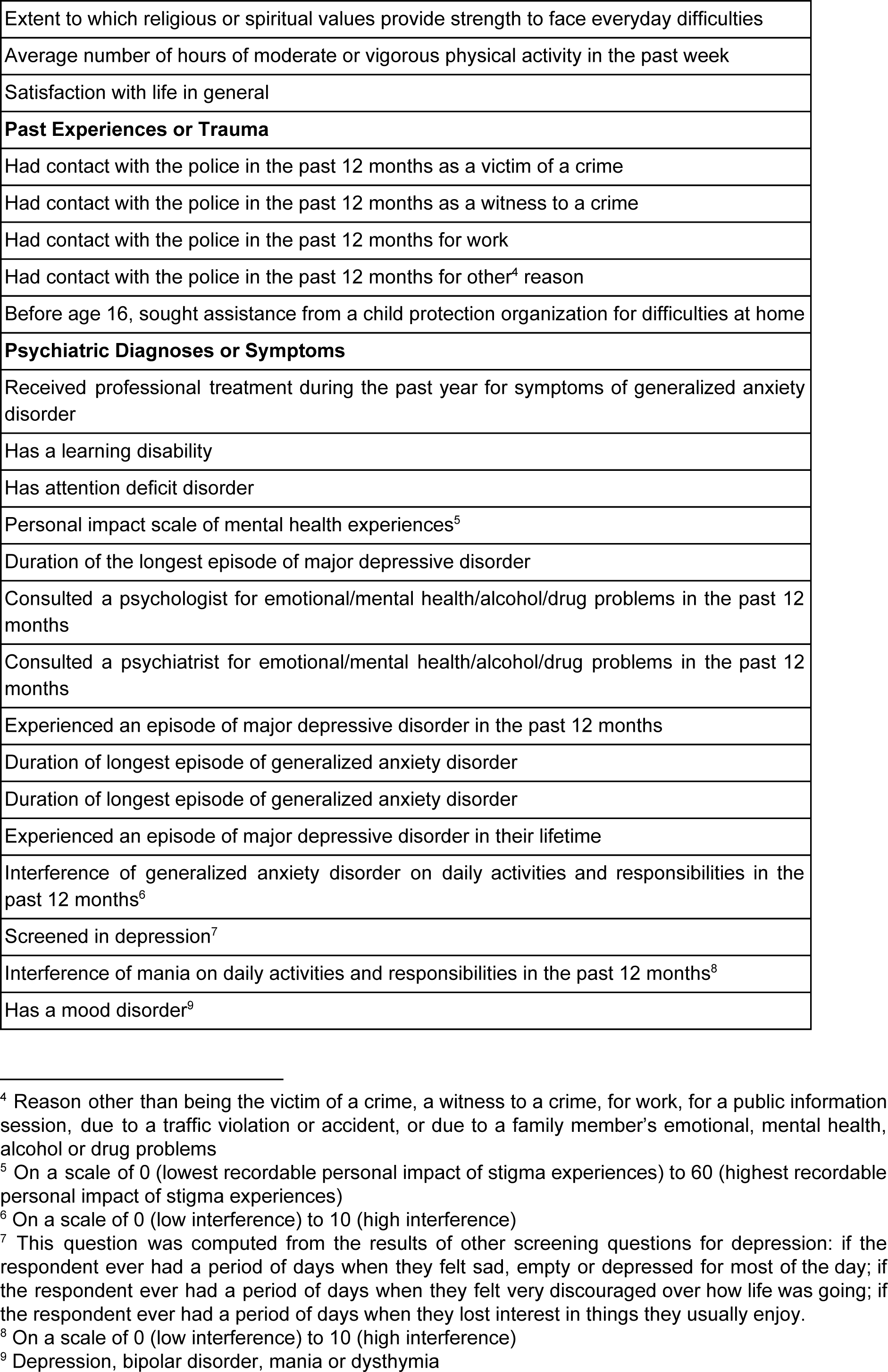
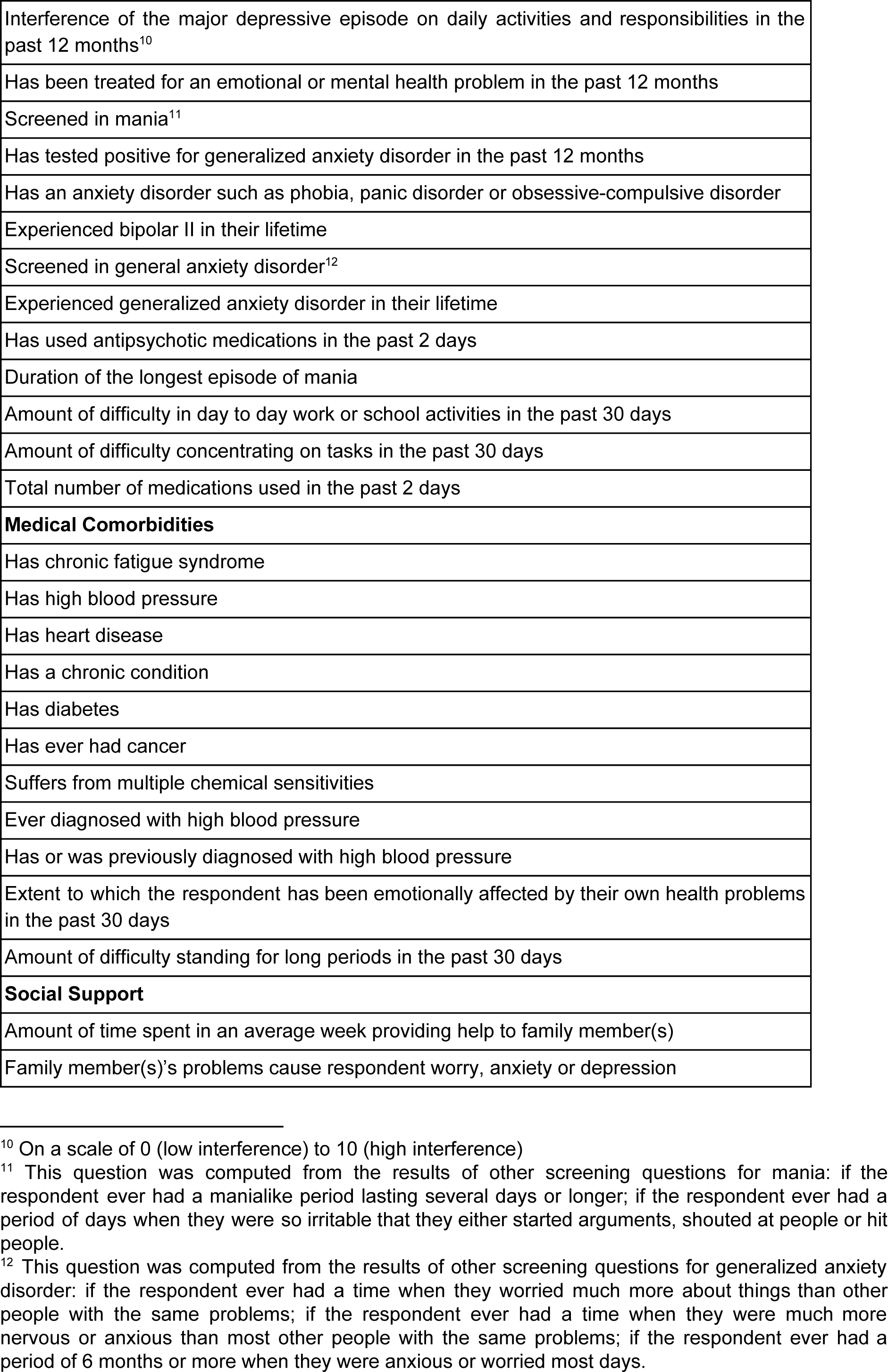
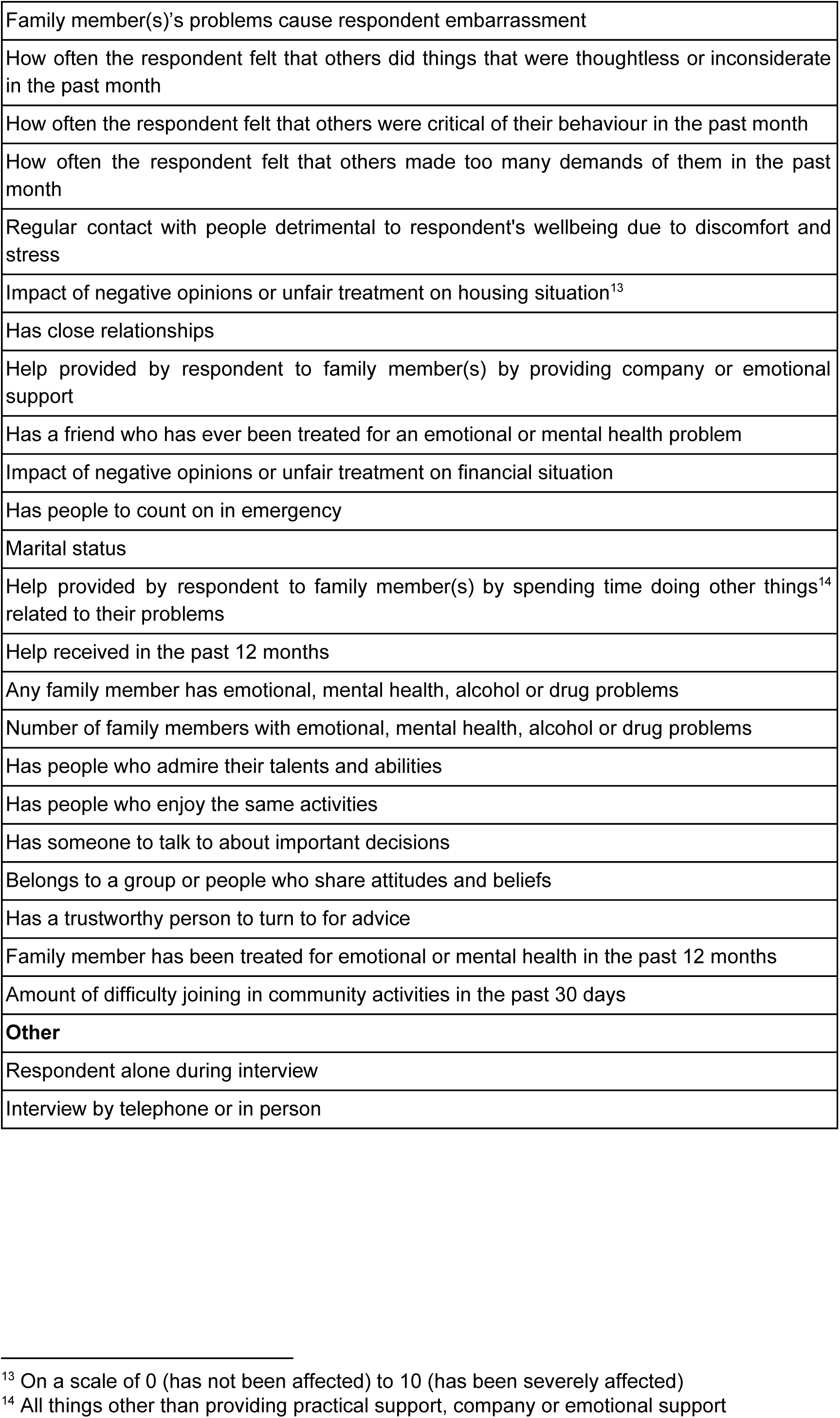
Features retained in the 96 feature version of the lifetime suicidal ideation prediction model

|  |
| --- |
| **Sociodemographic** |
| White or non-white race/visible minority |
| Occupation group |
| Currently pregnant |
| Currently attending school, college, CEGEP, or university |
| Time in Canada since immigration |
| Employment status last week |
| World Health Organization (WHO) Disability Assessment Schedule (WHODAS) score |
| **Lifestyle** |
| Number of cigarettes smoked per day (former daily smokers) |
| Type of smoker |
| Body Mass Index (BMI) (self-reported) |
| Experienced drug abuse or dependence (including cannabis) in their lifetime |
| Self-rated physical health |
| Experienced alcohol abuse or dependence in their lifetime |
| Experienced alcohol abuse in their lifetime |
| Type of smoker (calculated)<sup>3</sup> |
| Self-perceived rating of ability to handle day-to-day demands |
| Importance of religious or spiritual values in daily life |
| Number of years since stopped smoking daily (former daily smokers) |
| Self-perceived personal ability to deal with stress |
| Has ever smoked a whole cigarette |

<sup>3</sup> This question was computed from the results of other questions about smoking: if the respondent has smoked 100 or more cigarettes during their lifetime, if the respondent has ever smoked a whole cigarette, what type of smoker the respondent is, if the respondent has ever smoked cigarettes daily. This calculation produces a 1 (daily smoker) to 6 (never smoked) score.

|  |
| --- |
| Extent to which religious or spiritual values provide strength to face everyday difficulties |
| Average number of hours of moderate or vigorous physical activity in the past week |
| Satisfaction with life in general |
| **Past Experiences or Trauma** |
| Had contact with the police in the past 12 months as a victim of a crime |
| Had contact with the police in the past 12 months as a witness to a crime |
| Had contact with the police in the past 12 months for work |
| Had contact with the police in the past 12 months for other<sup>4</sup> reason |
| Before age 16, sought assistance from a child protection organization for difficulties at home |
| **Psychiatric Diagnoses or Symptoms** |
| Received professional treatment during the past year for symptoms of generalized anxiety disorder |
| Has a learning disability |
| Has attention deficit disorder |
| Personal impact scale of mental health experiences<sup>5</sup> |
| Duration of the longest episode of major depressive disorder |
| Consulted a psychologist for emotional/mental health/alcohol/drug problems in the past 12 months |
| Consulted a psychiatrist for emotional/mental health/alcohol/drug problems in the past 12 months |
| Experienced an episode of major depressive disorder in the past 12 months |
| Duration of longest episode of generalized anxiety disorder |
| Duration of longest episode of generalized anxiety disorder |
| Experienced an episode of major depressive disorder in their lifetime |
| Interference of generalized anxiety disorder on daily activities and responsibilities in the past 12 months<sup>6</sup> |
| Screened in depression<sup>7</sup> |
| Interference of mania on daily activities and responsibilities in the past 12 months<sup>8</sup> |
| Has a mood disorder<sup>9</sup> |

<sup>4</sup> Reason other than being the victim of a crime, a witness to a crime, for work, for a public information session, due to a traffic violation or accident, or due to a family member's emotional, mental health, alcohol or drug problems
<sup>5</sup> On a scale of 0 (lowest recordable personal impact of stigma experiences) to 60 (highest recordable personal impact of stigma experiences)
<sup>6</sup> On a scale of 0 (low interference) to 10 (high interference)
<sup>7</sup> This question was computed from the results of other screening questions for depression: if the respondent ever had a period of days when they felt sad, empty or depressed for most of the day; if the respondent ever had a period of days when they felt very discouraged over how life was going; if the respondent ever had a period of days when they lost interest in things they usually enjoy.
<sup>8</sup> On a scale of 0 (low interference) to 10 (high interference)

|  |
| --- |
| Interference of the major depressive episode on daily activities and responsibilities in the past 12 months<sup>10</sup> |
| Has been treated for an emotional or mental health problem in the past 12 months |
| Screened in mania<sup>11</sup> |
| Has tested positive for generalized anxiety disorder in the past 12 months |
| Has an anxiety disorder such as phobia, panic disorder or obsessive-compulsive disorder |
| Experienced bipolar II in their lifetime |
| Screened in general anxiety disorder<sup>12</sup> |
| Experienced generalized anxiety disorder in their lifetime |
| Has used antipsychotic medications in the past 2 days |
| Duration of the longest episode of mania |
| Amount of difficulty in day to day work or school activities in the past 30 days |
| Amount of difficulty concentrating on tasks in the past 30 days |
| Total number of medications used in the past 2 days |
| **Medical Comorbidities** |
| Has chronic fatigue syndrome |
| Has high blood pressure |
| Has heart disease |
| Has a chronic condition |
| Has diabetes |
| Has ever had cancer |
| Suffers from multiple chemical sensitivities |
| Ever diagnosed with high blood pressure |
| Has or was previously diagnosed with high blood pressure |
| Extent to which the respondent has been emotionally affected by their own health problems in the past 30 days |
| Amount of difficulty standing for long periods in the past 30 days |
| **Social Support** |
| Amount of time spent in an average week providing help to family member(s) |
| Family member(s)'s problems cause respondent worry, anxiety or depression |

<sup>10</sup> On a scale of 0 (low interference) to 10 (high interference)
<sup>11</sup> This question was computed from the results of other screening questions for mania: if the respondent ever had a manialike period lasting several days or longer; if the respondent ever had a period of days when they were so irritable that they either started arguments, shouted at people or hit people.
<sup>12</sup> This question was computed from the results of other screening questions for generalized anxiety disorder: if the respondent ever had a time when they worried much more about things than other people with the same problems; if the respondent ever had a time when they were much more nervous or anxious than most other people with the same problems; if the respondent ever had a period of 6 months or more when they were anxious or worried most days.

|  |
| --- |
| Family member(s)'s problems cause respondent embarrassment |
| How often the respondent felt that others did things that were thoughtless or inconsiderate in the past month |
| How often the respondent felt that others were critical of their behaviour in the past month |
| How often the respondent felt that others made too many demands of them in the past month |
| Regular contact with people detrimental to respondent's wellbeing due to discomfort and stress |
| Impact of negative opinions or unfair treatment on housing situation<sup>13</sup> |
| Has close relationships |
| Help provided by respondent to family member(s) by providing company or emotional support |
| Has a friend who has ever been treated for an emotional or mental health problem |
| Impact of negative opinions or unfair treatment on financial situation |
| Has people to count on in emergency |
| Marital status |
| Help provided by respondent to family member(s) by spending time doing other things<sup>14</sup> related to their problems |
| Help received in the past 12 months |
| Any family member has emotional, mental health, alcohol or drug problems |
| Number of family members with emotional, mental health, alcohol or drug problems |
| Has people who admire their talents and abilities |
| Has people who enjoy the same activities |
| Has someone to talk to about important decisions |
| Belongs to a group or people who share attitudes and beliefs |
| Has a trustworthy person to turn to for advice |
| Family member has been treated for emotional or mental health in the past 12 months |
| Amount of difficulty joining in community activities in the past 30 days |
| **Other** |
| Respondent alone during interview |
| Interview by telephone or in person |

<sup>13</sup> On a scale of 0 (has not been affected) to 10 (has been severely affected)
<sup>14</sup> All things other than providing practical support, company or emotional support

**Table 2.** Features retained in the 21 feature version of the lifetime suicidal ideation prediction model. Results of sensitivity analysis expressed as total numbers and ratios are presented in the middle and right columns. These express how many people would be classified as having suicidal ideation if all respondents tested gave answers at one or another extreme within the value range for a given question. For example, in row one, if all tested participants answered that they had stopped smoking less than 1 year ago, then there would be 6,819 positive classifications of suicidal ideation, and this would drop to 5,195 if all samples.had stopped smoking 3 or more years ago. The right most column describes the ratio of these two numbers.

| Feature Description | Model Sensitivity: Possible Response (number of samples classified as having suicidal ideation) | Model Sensitivity: Ratios |
| --- | --- | --- |
| **Sociodemographic** |  |  |
| **Lifestyle** |  |  |
| Opinion of own weight (self-reported) | Overweight (6,362), Just about right (4,799) | Overweight vs. Just about right (1.33 : 1) |
| When stopped smoking completely (former daily smokers) | Less than 1 year (6,819), 3 or more years (5,195) | Less than 1 year vs. 3 or more years: (1.31 : 1) |
| Has experienced alcohol abuse in their lifetime | Yes (6,452), No (5,220) | Yes vs. No (1.24 : 1) |
| Ever smoked cigarettes daily | Yes (6,608), No (6,160) | Yes vs. No (1.07 : 1) |
| Average number of hours of moderate or vigorous physical activity in the past week | 13.5 (5,998), 0 (5,144) | 13.5 hours vs. 0 hours of physical activity (1.06 : 1) |
| Used any medications for mental health, alcohol or drugs in the past 2 days | No (5,398), Yes (5,145) | No vs. Yes: (1.05 : 1) |
| Engaged in moderate or vigorous physical activity in the past week | Yes (5,607), No (5,334) | Yes vs. No (1.05 : 1) |
| **Past Experiences or Trauma** |  |  |
| Number of times before age 16 experienced sexual touching | More than 10 times (9,677), Never (4,790) | More than 10 times vs. Never: (2.02 : 1) |
| **Psychiatric Diagnoses or Symptoms** |  |  |
| Score on the Kessler Psychological Distress Scale (K6) in the past month | Highest Recordable Distress (19,758), Lowest Recordable Distress (1,545) | Highest vs. Lowest Recordable Distress (12.79 : 1) |
| Screened in generalized anxiety disorder | Yes (7,663), No (3,219) | Yes vs. No (2.38 : 1) |
| Personal impact scale of mental health experiences<sup>15</sup> | Lowest recordable personal impact of stigma experiences (15,520), Highest recordable personal impact of stigma experiences (8,229) | Lowest recordable personal impact of stigma experiences vs. Highest recordable personal impact of stigma experiences (1.89 : 1) |

<sup>15</sup> On a scale of 0 (lowest recordable personal impact of stigma experiences) to 60 (highest recordable personal impact of stigma experiences)

|  |  |  |
| --- | --- | --- |
| Has tested positive for generalized anxiety disorder in the past 12 months | No (5,391), Yes (4,016) | No vs. Yes (1.34 : 1) |
| Has post-traumatic stress disorder | Yes (6,983), No (5,384) | Yes vs. No (1.17 : 1) |
| Duration of the longest episode of major depressive disorder | 5 years or more (2,385), 2 weeks (2,052) | 5 years or more vs. 2 weeks (1.16 : 1) |
| Interference of depression on daily activities and responsibilities in the past 12 months | 0 (7,290), 9.6 (7,102) | Score of 0 vs score of 9.6 (1.02 : 1) |
| **Medical Comorbidities** |  |  |
| Has or was previously diagnosed with cancer | No (5,445), Yes (4,654) | No vs. Yes (1.16 : 1) |
| Has ever been diagnosed with high blood pressure | Yes (6,315), No (5,793) | Yes vs. No (1.09 : 1) |
| Has or was previously diagnosed with high blood pressure | Yes (5,424), No (5,397) | Yes vs. No (1.01 : 1) |
| **Social Support** |  |  |
| Has a trustworthy person to turn to for advice | Strongly disagree (9,350), Strongly agree (4,898) | Strongly disagree vs. strongly agree (1.91 : 1) |
| Has people to count on in emergency | Strongly disagree (8,046), Strongly agree (5,158) | Strongly disagree vs. strongly agree (1.56 : 1) |
| Impact of negative opinions or unfair treatment on housing situation | Has not been affected (18,631), Has been severely affected (17,508) | Has not been affected vs. has been severely affected (1.06 : 1) |

**Table 3.**
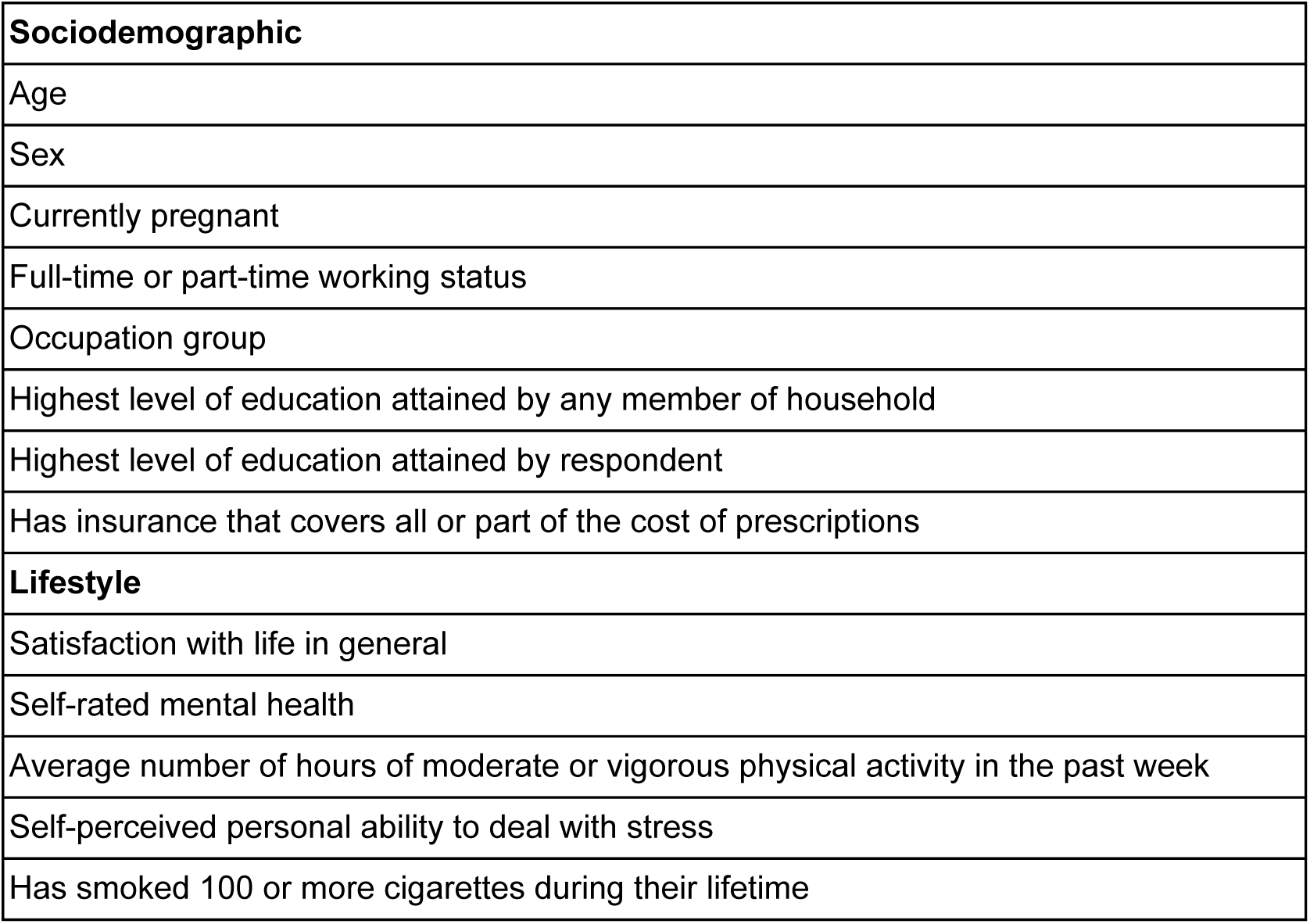
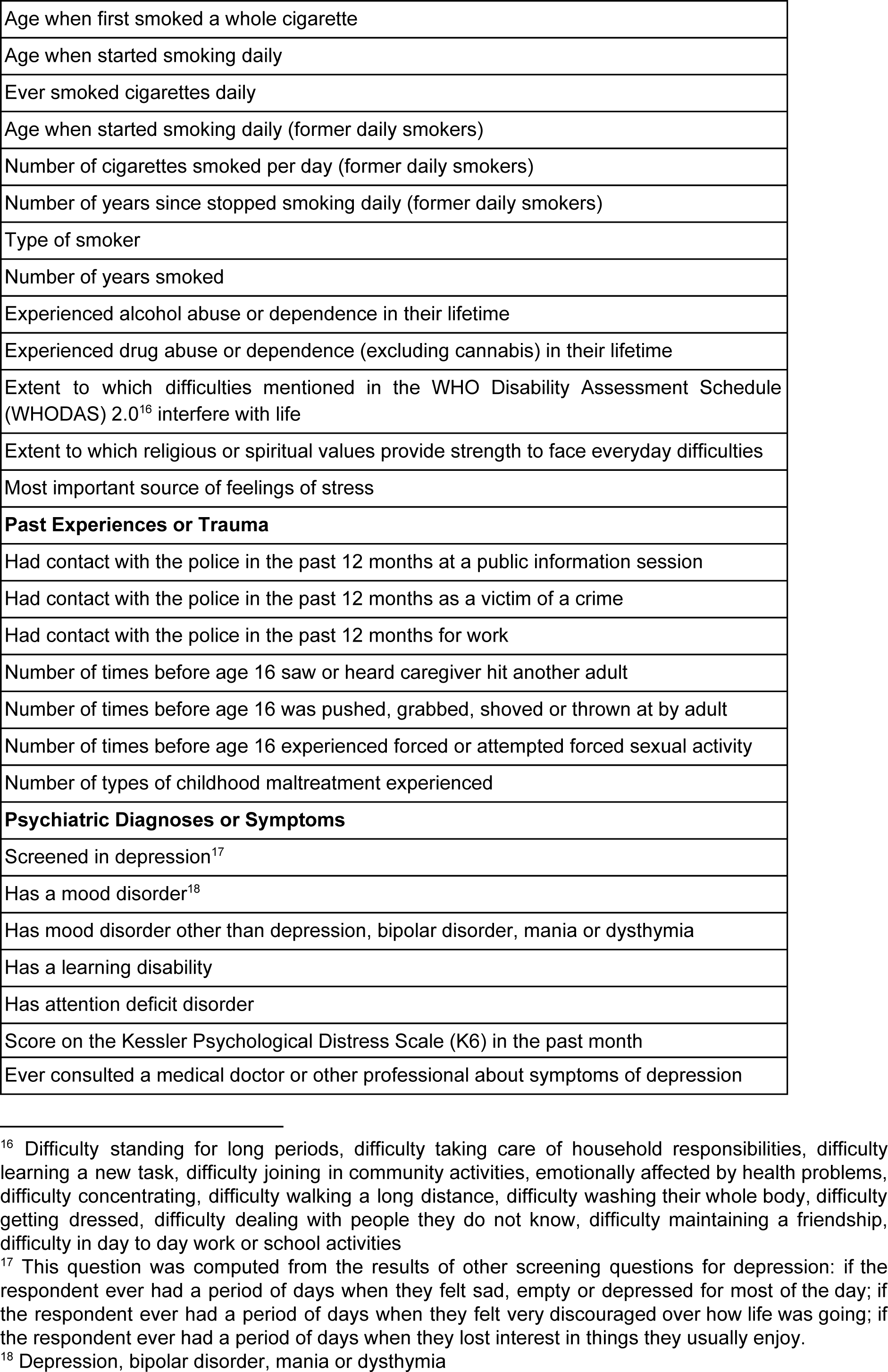
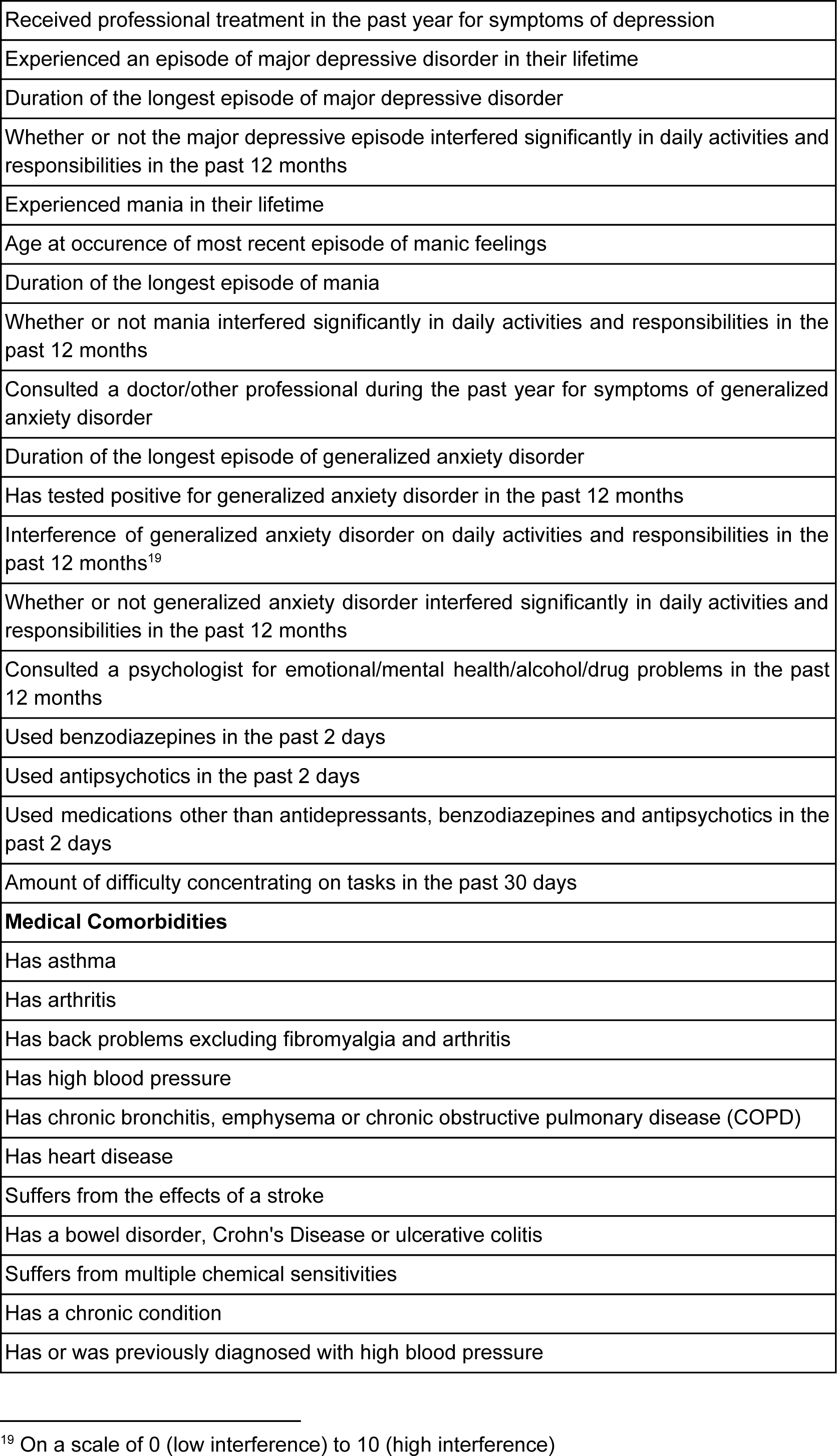
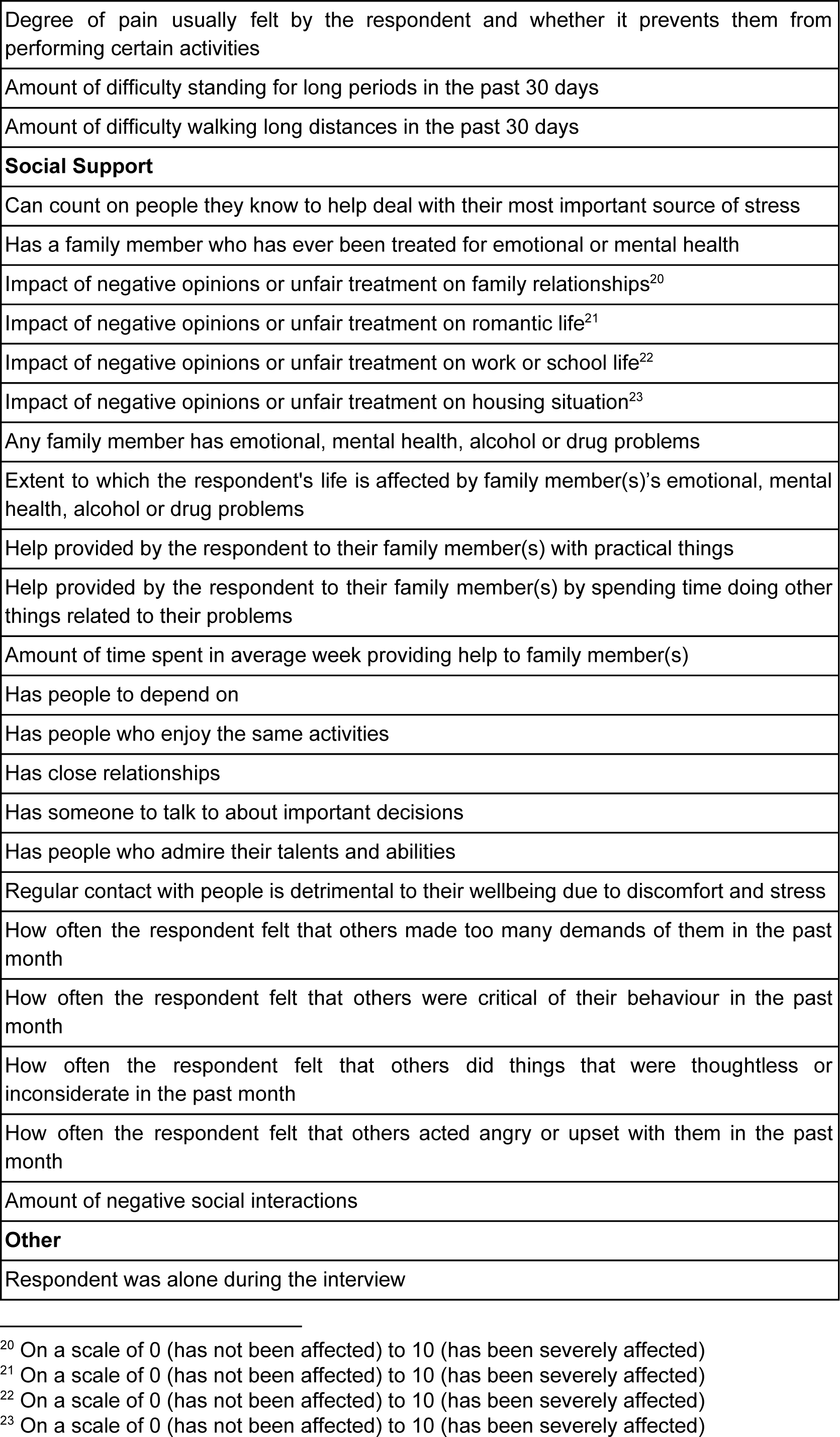
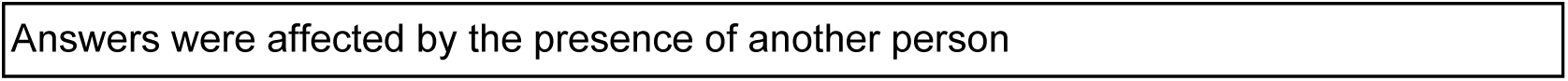
Features retained in the 96 feature version of the last 12 months suicidal ideation prediction model.

**Table 4.**
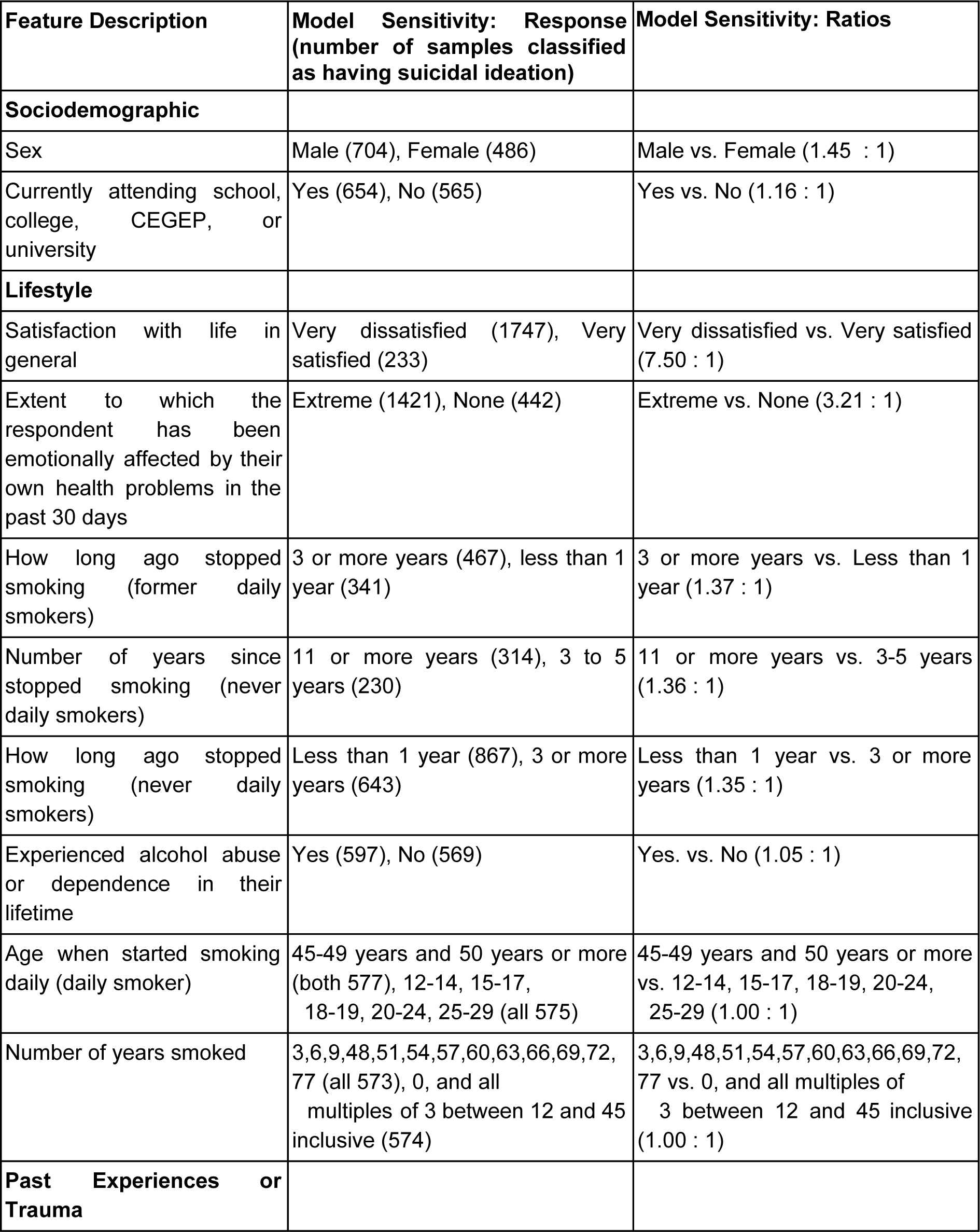
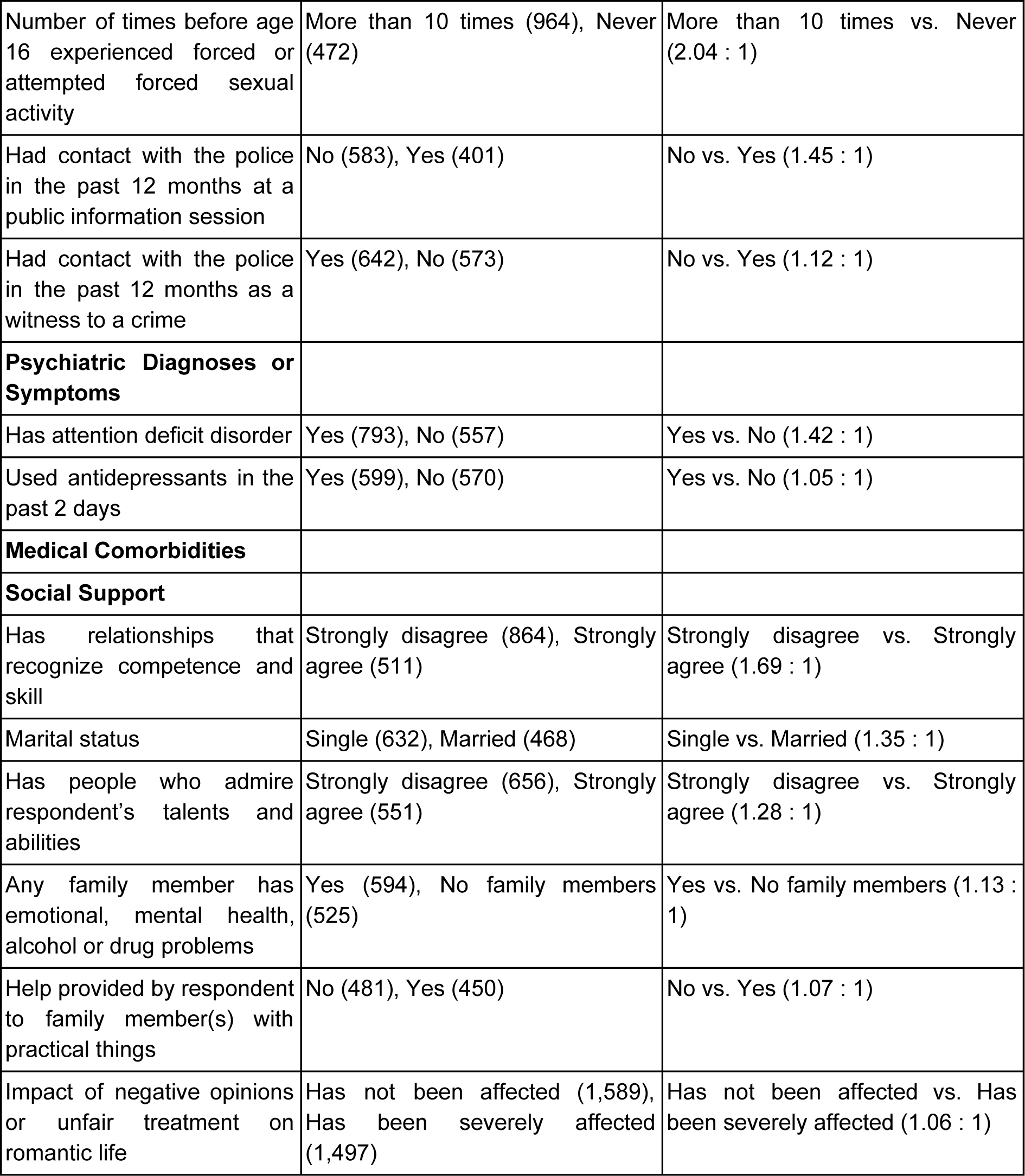
Features retained in the 21 feature version of the last 12 months suicidal ideation prediction model. Results of sensitivity analysis expressed as total numbers and ratios are presented in the middle and right columns.

**Table 5.**
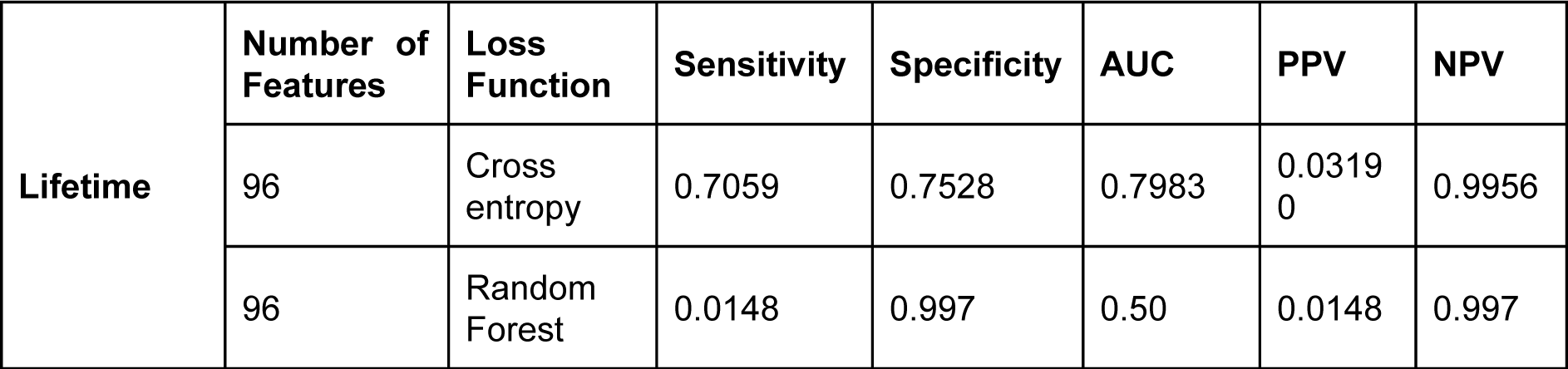
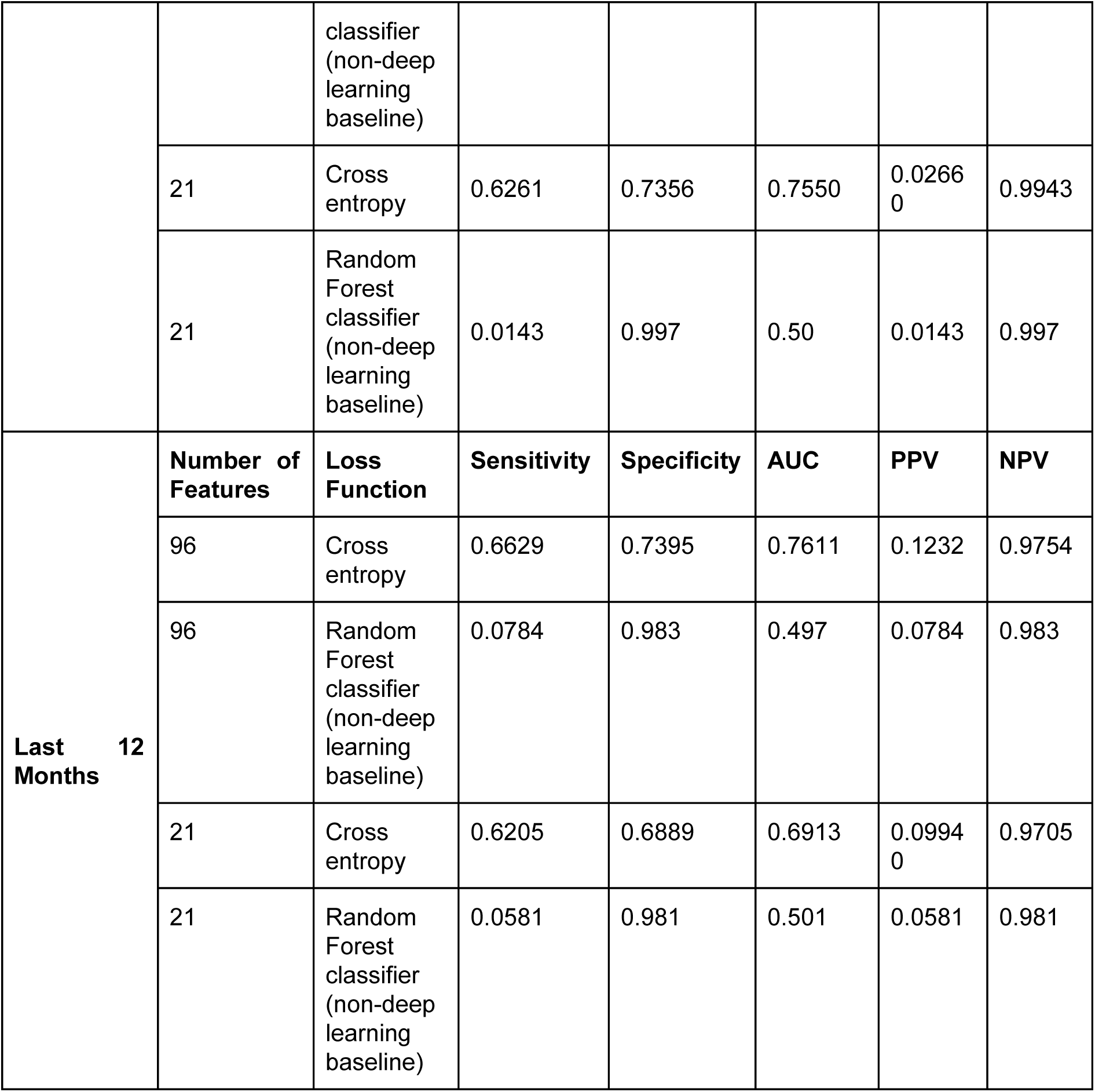
Lifetime and last 12 months suicidal ideation prediction model metrics, including comparison between random forest baseline model and deep learning (cross entropy loss function) results.

|  | **Number of Features** | **Loss Function** | **Sensitivity** | **Specificity** | **AUC** | **PPV** | **NPV** |
| --- | --- | --- | --- | --- | --- | --- | --- |
| **Lifetime** | 96 | Cross entropy | 0.7059 | 0.7528 | 0.7983 | 0.03190 | 0.9956 |
|  | 96 | Random Forest classifier<br>(non-deep<br>learning<br>baseline) | 0.0148 | 0.997 | 0.50 | 0.0148 | 0.997 |
|  | 21 | Cross<br>entropy | 0.6261 | 0.7356 | 0.7550 | 0.0266<br>0 | 0.9943 |
|  | 21 | Random<br>Forest<br>classifier<br>(non-deep<br>learning<br>baseline) | 0.0143 | 0.997 | 0.50 | 0.0143 | 0.997 |
| **Last 12**<br>**Months** | **Number of**<br>**Features** | **Loss**<br>**Function** | **Sensitivity** | **Specificity** | **AUC** | **PPV** | **NPV** |
|  | 96 | Cross<br>entropy | 0.6629 | 0.7395 | 0.7611 | 0.1232 | 0.9754 |
|  | 96 | Random<br>Forest<br>classifier<br>(non-deep<br>learning<br>baseline) | 0.0784 | 0.983 | 0.497 | 0.0784 | 0.983 |
|  | 21 | Cross<br>entropy | 0.6205 | 0.6889 | 0.6913 | 0.0994<br>0 | 0.9705 |
|  | 21 | Random<br>Forest<br>classifier<br>(non-deep<br>learning<br>baseline) | 0.0581 | 0.981 | 0.501 | 0.0581 | 0.981 |

**Table 7.** Sizes of datasets and distribution of data.

|  | Lifetime Suicidal Ideation | Last 12 Months Suicidal Ideation |
| --- | --- | --- |
| Total number of examples | 23,859 | 3,441 |
| Number of examples that answered 'yes' | 2,262 | 929 |
| Number of examples that answered 'no' | 21,597 | 2,512 |
| Number of males | 10,871 | 1,437 |
| Number of females | 12,988 | 2,004 |
| 60 TO 64 YEARS | 2,104 | 280 |
| 55 TO 59 YEARS | 2,097 | 376 |
| 15 TO 19 YEARS | 1,932 | 291 |
| 20 TO 24 YEARS | 1,886 | 330 |
| 65 TO 69 YEARS | 1,844 | 209 |
| 50 TO 54 YEARS | 1,821 | 329 |
| 30 TO 34 YEARS | 1,772 | 279 |
| 35 TO 39 YEARS | 1,616 | 279 |
| 40 TO 44 YEARS | 1,585 | 282 |
| 45 TO 49 YEARS | 1,571 | 254 |
| 80 YEARS OR MORE | 1,560 | 72 |
| 25 TO 29 YEARS | 1,521 | 279 |
| 70 TO 74 YEARS | 1,388 | 114 |
| 75 TO 79 YEARS | 1,162 | 67 |

### 2.2 Neural Network

The neural network used was a feed-forward fully-connected network with three hidden layers of 400 neurons each activated by the scaled exponential linear unit (SELU) function (Klambauer et al., 2017). SELU activation paired with AlphaDropout (Klambauer et al., 2017) maintains a self-normalizing property of the trained parameters of the network so as to keep the training procedure stable. Adam (Kingma & Ba, 2015) optimization was used to train the network. The final prediction layer had a softmax activation, allowing the network to establish its prediction in the form of a probability for both output classes.

### 2.3 Approach

In order to obtain a model that could be implemented in a real clinical environment, reducing the number of input features to pinpoint the most important features in the dataset was necessary-as a model that required too many input features would present challenges for data collection in the clinic when trying to apply the model to a given patient rapidly and efficiently^2^. The techniques used forfeature selection involved both expertise in the field (i.e. expert feature reduction) and allowing the model to highlight which features were the most important (Guyon & Elisseeff, 2003). A clinician (D.B.) went through all 582 features and discarded the features which were either administrative (i.e. redundant case identification codes or different ways of asking the same question) or which were not reasonable to collect clinically (such as detailed health care service satisfaction metrics which would not be appropriate in a screening context where the patient has not yet experienced services fully). This reduced the feature set size to 196.We further reduced the number of features using machine learning techniques. This involved analyzing the receptive fields of the trained model’s first layer and removing “unimportant” features. Feature “importance” was calculated via the weights that the neural network applied to a particular feature (Coates & Ng, 2011). Two cases were examined, one in which 100 features were removed, leaving 96 features in the model, and one in which 175 features were removed, leaving 21 features in the model. We chose to remove 100 and 175 features respectively, since the 100 feature removal didn’t affect the performance too much from the larger feature set sizes (> 100 features) and stopped at 175 because removing any more features would cause the performance to deteriorate vastly. The larger models were produced in order to maximize the identification of important features and to maximize model accuracy; the smaller models were produced in order to generate clinically tractable models with few enough questions that they could be integrated into a standard screening assessment. Separate models were produced for both lifetime and last 12 months suicidal ideation prediction.

In order to adjust our model to the large class imbalance that existed between the “no” and “yes” responders, we used undersampling. The number of examples in the majority (“no”) class was equated to the number in the minority (“yes”) class. In the case of lifetime prediction, 2,262 random examples from the “no” class were randomly chosen for the training set to match the 2,262 samples from the “yes” class. The class-balanced training set was then divided into 10 different random folds, and the model was trained on 9 of these folds, leaving the final fold and all of the other 19,355 “no’s” to serve as the validation set. This process was repeated 10 times with mutually exclusive validation and training sets, and we noted the average of the test metrics of all runs on the validation set. It is important to mention here that our validation set comprised of a relatively lower count of respondents in the “yes” class compared to the initial distribution of the data, making it much harder for the model to be able to classify respondents in the “yes” class correctly. The same sort of division was performed for the last 12 months data using the data distribution shown in Table 7.

Given that classifying an individual to *not* have suicidal ideation when they actually are experiencing suicidal ideation is a more costly error than predicting the inverse, we penalized false negatives harder than other classifications. Penalization was achieved by summing to the cost function the penalty factor defined by the number of false negatives to the power of five.

All analyses were done using the Vulcan software package (see software note). Figure 1 represents the steps taken to produce the results for this analysis.

**Figure 1.**
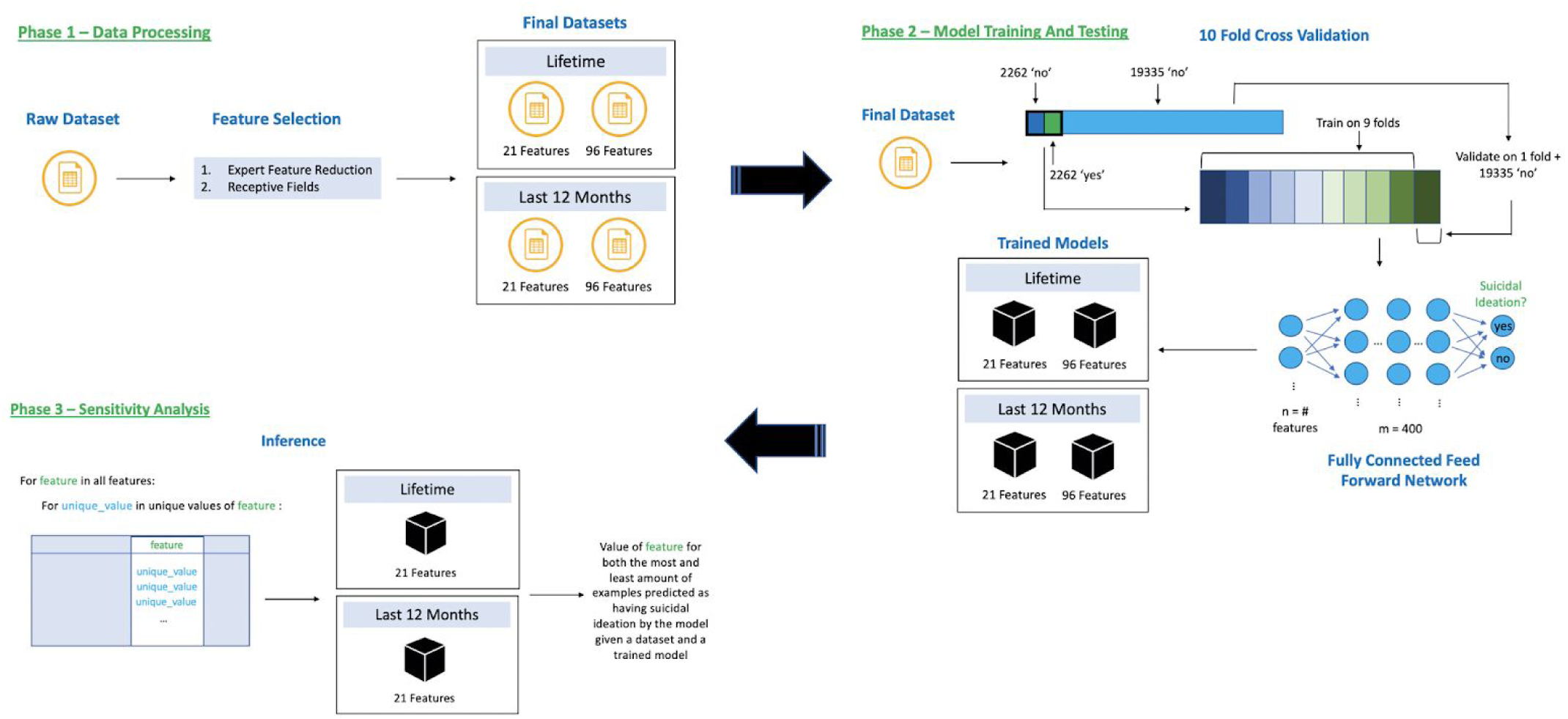
Flow of data through our training and inference system broken into three phases, 1) Data Processing – reduce dataset features using expert reduction & receptive fields, 2) Model Training & Testing – 10-fold cross validation using under sampling of the ‘yes’ class and training a neural network, and 3) Sensitivity Analysis – discovering feature directionality for our 21 feature trained models.

## Results

Tables 1-4 show the features used for the prediction of lifetime suicide ideation (Table 1: 96 features, Table 2: 21 features) and suicide ideation during the past 12 months (Table 3: 96 features, Table 4: 21 features). These features are those that remain following expert feature reduction (manual feature removal using domain expertise) and using the network’s first layer receptive fields to remove additional features until 96 and 21 features remained for both lifetime and last 12 months suicidal ideation models. In terms of measurement, we chose to use the AUC (area under the receiver operating curve) as our main metric of model performance, and we also calculated the sensitivity, specificity, negative predictive value (NPV) and positive predictive value (PPV) for each model. Tables 5 and 6 show the 10-fold cross validated results for the lifetime (96 features - 0.7983 AUC; 21 features - 0.7550 AUC) and last 12 months (96 features - 0.7611 AUC; 21 features - 0.6913 AUC) datasets, respectively. Random forest classifiers were produced as a non-deep learning baseline; these generally performed poorly in terms of AUC (Tables 5 and 6). In total, we produced four model configurations : 96 and 21 features for predicting lifetime suicidal ideation and 96 and 21 features for predicting suicidal ideation in the last 12 months.

In order to gain insight into how different features affected model predictions (i.e. feature directionality), we performed a feature sensitivity analysis for the 21 feature models. We chose not to perform the same analysis for the 96 feature models as it would be unsuitable to interpret due to size. We explored how variations in values for a specific feature affected the final model prediction. We accomplished this by iterating through all possible unique values (up to a maximum of 20 values) for each feature and imputed all response samples to have this value. We then ran a test to determine how many of the samples would be classified as having suicidal ideation by the model. The rightmost columns in the 21 feature tables (Tables 2, 4) show the value of the feature where the model predicted the most amount of suicidal ideation followed by the feature value with the lowest amount of suicidal ideation. In Tables 2 and 4, the number in brackets next to each feature value shows the number of examples in the test set classified as having suicidal ideation (19,788 samples in the test set for lifetime; 1,769 for the past 12 months). This allows for some insight into the inner workings of the neural network model. For example, in the lifetime prediction of suicidal ideation, if all the answers to the question “have people to count on in an emergency” are set to “strongly disagree”, then 8,046 people are predicted to have suicidal ideation; this number drops to 5,158 people if the answers are all changed to “strongly agree”.

## Discussion

Here we illustrate that using our method, suicidal ideation data from the general populattion can identify people at high risk for suicide, who could likely benefit from more in-depth screening and resources in the context of suicide prevention.

Jordan et al. (2018) found that using only four items of the PHQ-9 provided the most accurate predictions of suicidal ideation in their patient sample - those assessing “feelings of depression/hopelessness, low self-esteem, worrying, and severe sleep disturbances” (Jordan et al., 2018). Although the PHQ-9 was not included in our dataset, our model similarly found some high impact variables related to depression, hopelessness and worrying. For instance, a high score on the Kessler Psychological Distress Scale (K6), which assesses feelings of depression and hopelessness, seems to be a significant risk factor for suicidal ideation (Tables 2 and 3). Unlike the Jordan model, ours did not identify sleep problems to be a significant risk factor for suicidal ideation. One possible explanation accommodating our results and those in the literature is that sleep problems, rather than being a risk factor themselves, may act as a proxy for actual interacting risk factors. When such factors are included in the data and processed by a complex model, sleep disorder factors are rendered “irrelevant”. We will seek to verify this hypothesis in other datasets with more robust measures of sleep. While Jordan’s model identified low self-esteem as a risk factor, our dataset unfortunately did not contain a self-esteem variable. Our model yielded additional predictive factors that do not overlap with those found by the Jordan team. Generalized anxiety disorder, for example, appears to be an important predictor of suicidal ideation (Tables 1, 2, and 3). This is to be expected, since previous research has identified anxiety disorders, including generalized anxiety disorder, as independently predictive of suicidal ideation (Bentley et al., 2016; Sareen et al., 2005). Importantly, our method yielded predictors related to early sexual experiences and sexual abuse. Sexual experiences before the age of 16, including non-consensual experiences, appear to be important risk factors for suicidal ideation (Tables 2, 3 and 4). This finding is supported by previous research linking increased suicidal ideation and suicide attempts to early sexual abuse (Basile et al., 2006, Bedi et al., 2011, Lopez-Castroman et al., 2013, Thompson et al., 2018, Ullman et al., 2009), thus confirming our model’s capacity to identify known risk factors of suicidal ideation. Childhood sexual abuse is a particularly important consideration in suicide prevention. There is extensive literature suggesting that early-life adversity, including childhood sexual abuse, is an important predictor of suicidal behavior (Turecki & Brent, 2016; Wanner et al., 2012; Brezo et al., 2008).

We separated prediction of suicidal ideation occuring in the last 12 months and throughout the lifetime to disambiguate more specific short term from long term predictors. Identification of protective factors and risk factors for both conditions may improve methods of identifying and treating those at risk of attempting suicide. Lifetime factors may be useful in developing more long term suicide prevention strategies, while factors predicting suicide ideation in the last 12 months can inform the identification and treatment of patients at more immediate risk. While all predictors were related to wellbeing, mental health, early sexual experiences and sexual abuse, we found important differences between risk factors and protective factors for the lifetime and last 12 months conditions. Features related to social support, such as marital status and having relationships that recognize competence and skill, seem to be more influential in predicting suicidal ideation in the past year than throughout the lifetime. This may indicate that measures of social support could be used to identify patients at more immediate risk of suicidal ideation. Based on previous literature, lack of social support may be a moderator between life stress and suicidal ideation, suggesting that a strong social support system may be beneficial in reducing suicidal thoughts particularly during stressful times (Vanderhorst & Dr, 2005; Yang & Clum, 1994). Additionally, the level to which one has been affected by their health problems in the previous 30 days, and dissatisfaction with life in general may be specific risk factors for suicidal ideation in the past 12 months. Both of these measures could be included as screening questions to identify patients who may be experiencing suicidal thoughts. By contrast, physical and mental health related features may have more long term effects on suicidal ideation because more health related features appear in the model predicting lifetime occurrences. As noted, depression and anxiety symptoms were important predictors of lifetime suicidal ideation. This may also be related to the timing of the data collection, as a smaller number of respondents would have been experienced a mood episode or high levels of anxiety during the interview year itself than over the course of their lifetimes.

We identified several surprising features that did not show a clear directionality in our sensitivity analysis. While these features are not clear risk or protective factors, they seem to interact with other features to predict suicidal ideation. Notably, features related to smoking, including when patients started and stopped smoking, appeared in both the lifetime and past 12 months prediction models as potential moderators of suicidal ideation. Previous explorations found that cigarette use increases the risk of suicidal ideation, a relationship that could potentially be explained by the lower levels of serotonin found in smokers (Malone et al., 2003; Tanskanen, Viinamäki, Hintikka, Koivumaa-Honkanen, & Lehtonen, 1998). Our results complement these findings, while alluding to a more complex relationship without clear directionality when other factors are considered. This supports a focus on public health and public mental health interventions on smoking. Additionally, contact with the police may moderate suicidal ideation, highlighting a need to follow up with people who may have had a traumatic experience leading to police intervention, or negative interactions with the police (DeVylder et al., 2018).

We identified several predictors that are easy to obtain, including sociodemographic features. Interestingly, Jordan et al. did not find sociodemographic features useful in the prediction of suicidal ideation (2018), but as we were using a census dataset with a large and varied array of sociodemographic features, we were able to identify more predictors amongst them that would be amenable to upstream intervention. As opposed to more expensive data like neuroimaging and genetic testing, sociodemographic predictors can be very useful in clinical practice, especially with respect to screening, since they are easily accessible to healthcare professionals through direct questioning or self-report questionnaires.

As can be seen in Tables 5 and 6, the 96 feature models have higher AUCs. This is expected, as the network is able to make better predictions when it has more information of the different patients it is classifying. It is worthwhile to discuss the pros and cons of having larger or smaller models. Large models which do not overfit allow us to identify more predictors which may be modifiable and are therefore potentially useful from a public health standpoint. Smaller models are easier to implement because patients need to answer fewer questions in order to provide the model with sufficient information to make a prediction. Thus, there exists an interesting trade-off between model accuracy and ease of data acquisition upon selecting the number of features to include.. For example, the difference in the AUC for the last 12 months model presented here is 0.69 for the 21 feature model vs. 0.76 for the 96 feature model. Does this 7 point difference justify a larger model that is more accurate but more difficult to collect? While 7 points may only seem like a moderate difference, when considering predictions on a population scale we might expect a significant difference in the absolute number of people correctly classified. Implementation of models such as these will hinge on finding the right balance between model complexity and accuracy in order to provide models that are both meaningful and feasible to implement.

It is also important to note the high negative predictive values (NPV) of our predictions. This metric indicates that the network is almost always correct when it classifies an example as not having suicidal ideation. This is crucial, as it signifies the utility of our model in helping, alongside good clinical judgement and history taking, to rule out suicidal ideation in populations matching those in the dataset. Given that currently, clinicians have difficulty ruling out suicidal thinking or risk (McDowell et al., 2011), such a tool would be clinically useful. This must be balanced against the risk of false positives, which can lead to unnecessary intervention and confinement, as well as against the fact that the absence of suicidal ideation at a single point in time does not rule out the risk of suicide (McHugh et al., 2019). However, given that this model predicts suicidal ideation and not risk of attempt, a positive result could be used to open a conversation between a clinician and patient, which might lead to more appropriate assessment and treatment before the risk of an attempt increases. This in turn may become a useful approach for the prevention of suicide via upstream identification of at-risk patients in the general population, though this remains speculative and should be expanded on in future work exploring factors that predict conversion of ideation to action.

There are several limitations to our current work. While using an interview-based census dataset allows for a large sample size in the general population, it does mean that there is no independent verification of participant responses or any clinician-rated scales. Our use of deep learning provides for a powerful technique that outperforms random forest classifiers, but which is generally less easy to interpret than other machine learning techniques; that being said, our sensitivity analysis does allow some insight into the model parameters which could be further evaluated using classical statistics.

It is worth discussing the practical implementation of a predictive tool of suicidal ideation in clinical practice, as this would bring both possible benefits and challenges. One possible implementation of this tool would be as an automated screening tool integrated into electronic medical records in emergency departments or outpatient clinics. Benefits - which would need to be verified in clinical studies - could include earlier and more accurate identification of suicidal ideation, which would lead to more patients being offered appropriate services, such as access to a therapist or to crisis resources. This in turn would hopefully lead to a reduction in the number of patients making suicide attempts or completing suicide, though this would depend on the efficacy of the offered interventions. Nonetheless, challenges and potential dangers exist. Models that predict suicidal ideation could be used by some clinicians to justify interventions such as forced hospitalization, which raises serious concerns about the effect of implementing such models on patient autonomy and clinician medico-legal risk. In addition, it is unclear what effect having an automated screening tool for suicidal ideation would have on clinician behavior. It might improve clinician awareness of the importance of screening for and offering support to patients with suicidal ideation; at the same time, it may reinforce the habit of many clinicians to avoid asking about suicidal ideation, fostering an over-reliance on an imperfect system to screen for a potentially serious clinical phenomenon. Any implementation of such a screening system would require significant investment in the training of clinicians and should be accomplished in partnership with patient and clinician representatives.

## Data Availability

Data available from Statistics Canada

## Supplementary Materials

**Table 8.** Sensitive cost function experiment results. The rows in Table 8 have the sensitive cost function that indicate an experimental approach where instead of using the simple cross entropy loss function, we added an extra penalty to the false negatives, in the hope of improving the number of false negatives. As can be seen, the sensitivity did improve slightly for the 96 and 21 feature lifetime datasets. However, it actually decreased the sensitivity in both data subsets for the last 12 months prediction. This could have been because the number of samples in the last 12 months dataset is extremely small, and thus the sensitive cost function did not have the desired effect of being exposed to suboptimal levels of data variation

| **Number of Features** | **Sensitivity** | **Specificity** | **AUC** | **PPV** | **NPV** |
| --- | --- | --- | --- | --- | --- |
| Lifetime - 96 | 0.7140 | 0.7436 | 0.7995 | 0.03120 | 0.9955 |
| Lifetime - 21 | 0.6369 | 0.7189 | 0.7486 | 0.02560 | 0.9941 |
| Last 12 months - 96 | 0.6603 | 0.7155 | 0.7494 | 0.1138 | 0.9744 |
| Last 12 months - 21 | 0.5909 | 0.6879 | 0.6884 | 0.0946 | 0.9683 |

### Methods Addendum

#### Variance Thresholding

Variance thresholding, a method which removes columns (i.e. features) if they do not vary sufficiently across the patient samples depending on the threshold given, was attempted but discontinued as it seemed to adversely affect the predictive power of the results. We presume this may have been due to the extreme imbalance in the dataset, where columns removed via this method may have in fact been the determining features that helped distinguish the difference between having suicidal ideation and not having them.

## Footnotes

2 A note on terminology: “feature” here refers to an input variable (i.e. one survey item).

